# The epidemiological impact of the Canadian COVID Alert App

**DOI:** 10.1101/2022.01.04.21268588

**Authors:** Shuo Sun, Mairead Shaw, Erica EM Moodie, Derek Ruths

## Abstract

**Objectives:** We analyzed the effectiveness of the Canadian COVID Alert app on reducing COVID-19 infections and deaths due to the COVID-19 virus.

**Methods:** Two separate, but complementary approaches, were taken. First, we undertook a comparative study to assess how the adoption and usage of the COVID Alert app compared to those of similar apps deployed in other regions. Next, we used data from the COVID Alert server and a range of plausible parameter values to estimate the numbers of infections and deaths averted in Canada using a model that combines information on number of notifications, secondary attack rate, expected fraction of transmissions that could be prevented, quarantine effectiveness, and expected size of the full transmission chain in the absence of exposure notification.

**Results:** The comparative analysis revealed that the COVID Alert app had among the lowest adoption levels among apps that reported usage. Our model indicates that use of the COVID Alert app averted between 6,284 and 10,894 infections across the six Canadian provinces where app usage was highest during the March - July 2021 period. This range is equivalent to 1.6%-2.9% of the total recorded infections across Canada in that time. Using province-specific case fatality rates, 57-101 deaths were averted during the same period. The number of cases and deaths averted was greatest in Ontario, whereas the proportion of cases and death averted was greatest in Newfoundland and Labrador. App impact measures were reported so rarely and so inconsistently by other countries that the relative assessment of impact is inconclusive.

**Conclusions:** While the nationwide rates are low, provinces with widespread adoption of the app showed high ratios of averted cases and deaths (upper bound was greater than 60% of averted cases). This finding suggests that the COVID Alert app, when adopted at sufficient levels, can be an effective public health tool for combatting a pandemic such as COVID-19.

## 1. Introduction

The coronavirus disease (COVID-19) pandemic represents a substantial challenge for public health, pandemic planning, and health care systems. The pathogen, severe acute respiratory syndrome coronavirus 2 (SARS-CoV-2), is highly transmissible. Multiple public health strategies have been employed since the onset of the pandemic to reduce transmissions. Even with the widespread adoption of vaccines, many of these strategies remain in place, including contact tracing. In an effort to facilitate rapid identification and notification of exposure, several countries, including Canada, developed and adopted a smartphone-based exposure notification app. The Canadian app, COVID Alert, is the focus of our analysis. The main function of the app is exposure notification, which serves as a form of digital contact tracing. Any two phones that have the COVID Alert app downloaded and are near one another exchange random codes every 5 minutes using Bluetooth. If an app user receives a positive test result for COVID-19, they may be given a “one-time key” to enter into the app. All app users who have been in close contact with a user who entered a one-time key are sent a notification of exposure through the app. Close contact is defined as two phones being within 2 metres of one another (distance is estimated using the strength of the Bluetooth signal) for at least 15 minutes during the 14 days preceding the date that the positive test result is uploaded via the one-time key. The COVID Alert app was designed to offer a high degree of privacy to users and does not collect information on the name or address of the app user or the location of the user at any time (in particular, GPS information is not accessed or collected).

COVID Alert was piloted in the province of Ontario in July 2020 and more widely deployed in other provinces and territories of Canada in the fall of 2020. Newfoundland and Labrador, New Brunswick, and Saskatchewan adopted the app in September, followed by Manitoba, Quebec, Prince Edward Island, and Nova Scotia in October, and the Northwest Territories in November (COVID-19 Exposure Notification App Advisory Council, 2021). To date, the COVID Alert app has been downloaded on more than 6 million devices in the nine onboard provinces and territories. Roughly 71,000 notifications were sent as a result of 34,000 app users uploading one-time keys over the period from February 25, 2021 to July 18, 2021.

To investigate the epidemiological impact of the COVID Alert app on mitigating virus transmission in Canada, we undertook a survey of COVID contact notification apps in other regions and used epidemiological data from the Canadian COVID Alert app to estimate the number of cases and deaths averted due to the app during our study period of March to July, 2021 (Canadian Digital Service, n.d.). This article is organized as follows. In Section 2, we detail the methods for the search strategy for our comparative analysis and provide an overview of the modelling approach, with full details of the model provided in the Web Supplement. Section 3 presents the results of our analyses, and Section 4 discusses and concludes.

## 2. Methods

### 2.1 Data sources

The statistics used for this analysis were drawn from official, aggregate COVID Alert server and app daily usage statistics provided by the Canadian Digital Service (Canadian Digital Service, n.d.). At the time of printing, this dataset has not yet been released publicly, though Health Canada has plans to do so. To learn about or request access to the data, email Health Canada at.

Although the app was fully deployed by December 2020, it was not until a February 2021 update that many of the statistics required for modeling uptake and impact could be collected. Thus, our analysis considers the time period beginning February 25, 2021.

### 2.1 Comparative assessment

To compare the uptake and usage of COVID Alert to similar apps deployed in other countries, we first sourced countries with apps from a Wikipedia list of 47 countries with official exposure notification apps (COVID-19 apps, n.d.). We conducted a search for reports related to app uptake and efficacy first using Google Scholar, then regular Google search results and news sources. We used a snowball search strategy, collecting articles and news stories referenced in sources already gathered. Non-government sources were of variable quality; the source and timeline of reported information was often unclear and there was little consistency between news reports. In an effort to preserve the quality of comparison, we thus prioritized sources with direct access to app data, either government data/reports or research reports from teams working with app data directly. Of the 47 regions (countries or states) with apps, we were able to find 8 with direct access sources: France (TousAntiCovid;France, 2021; Cédric, 2021), Germany (Corona-Warn-App; Hoerdt, 2021), Italy (Immuni; Presidenza del Consiglio dei Ministri, n.d.), the Netherlands (Corona Melder; Boncz, 2021), New Zealand (NZ COVID Tracker; Ministry of Health NZ, n.d.), Switzerland (SwissCovid; Salathé et al., 2020), the United Kingdom (NHS COVID-19 app; Wymant et al., 2021) and USA’s Washington state (WA Notify; Segal et al., 2021).

We compared COVID Alert to the selected apps deployed in other countries or, in one case, the state of Washington in the USA along five metrics. To compare adoption and usage, we extracted: (1) app downloads, (2) active users, and (3) exposure notifications sent. To compare with the results of our modelling, we additionally extracted: (4) estimated cases averted and (5) estimated deaths averted. To facilitate fair comparisons, we considered these metrics by percentage of the regions’ population or by percentage of their total cases. Country populations and total cases were taken from Worldometer on July 27, 2021. The population and total case numbers for Washington state were drawn from the United States Census Bureau and the New York Times, respectively (Allen et al., n.d.; U.S. Census Bureau, n.d.).

### 2.2 Modeling

To assess the impact of COVID Alert on mitigating virus transmission, we estimated the number of COVID-19 cases averted in each province based on a modelling approach proposed by Wymant et al. (2021). In brief, these authors used an approach that models the number of cases averted due to notifications received on day *t* as the product of five terms: (i) the number of notifications received on day *t*, (ii) the secondary attack rate (SAR), which is the probability that someone who is notified will test positive, (iii) the expected fraction of transmissions preventable if an infectious individual strictly adheres to quarantine after receiving a notification, (iv) the quarantine effectiveness, and (v) the expected size of the full transmission chain that would originate from the contact if they had not been notified. Details of each of these quantities are provided in the Web Appendix.

As noted above, the COVID Alert app was rolled out at different dates in different provinces beginning in Ontario in July 2020. As mentioned earlier, as the statistics needed for modeling were only available as of February 25, 2021, our analysis considers the time period beginning on this date.

The process to obtain a one-time key to upload a positive COVID-19 test is different in each province and territory, thus it is possible that some users do not ever receive a key. Further, COVID-19 positive declaration is not mandatory and users who test positive have only 24 hours to enter the key in the app. Therefore, the number of notifications received during the study period is an underestimate of the number of users who test positive. Due to the constraints of privacy preservation, the SAR, the expected fraction of transmissions prevented, and the quarantine effectiveness cannot be estimated from the available data. We consider instead a range of plausible values for these parameters that are based on the literature (see Wymant et al., 2021 and Segal et al., 2021). In particular, we consider SARs of 5% and 6%. Following the results in Ferretti et al. (2020), the generation time (i.e., the time from infection of the index case to the time of infection of the secondary case) is modelled by a Weibull distribution with an average generation time of 5.5 days. The fraction of transmissions prevented is estimated from the delay distribution using the generation time distribution assuming that the mean time from exposure to notification among those app users is 5.46 days (as in Segal et al., 2021); this correlates to approximately 50% of transmissions being prevented by receipt of exposure notifications. For the effectiveness of quarantine in reducing transmission, two plausible values, 45% and 65%, were used (Wymant et al., 2021; Segal et al., 2021).

The size of the transmission chain is a function of the number of cases during the study period. It describes the number of cases at time *T* that are caused by transmissions originating from the contact if not notified before time *T*. Here we follow the assumptions of Wymant et al., 2021; specifically, it is assumed that local epidemics do not mix and that the extra cases do not affect the epidemic dynamic (i.e., the underlying epidemic growth rate does not change with the additional cases). We estimated the number of deaths averted by multiplying the number of cases averted by the province-specific crude case fatality rate, which was estimated for each province as the ratio of its total number of deaths due to the COVID-19 virus during the modeling time period to its total number of cases during the modeling time period. Note that these rates are a lower bound because the time delay from illness onset to death leads to right censoring; that is, the true number of deaths among those cases will be equal to or greater than the observed number at the end of the study because some people may die subsequent to the period of study.

## 3. Results

### 3.1 Comparative assessment of adoption and usage

The numbers of downloads and active users are summarized in Table 1, along with the periods of reporting. To facilitate the comparison between apps, both downloads and active users are also presented as a percentage of the total country or state population, and the number of active users is presented as a percentage of downloads. We were surprised to discover how few reliable usage and impact statistics were published for the many apps developed and deployed internationally. This presented an obstacle to any in depth comparative analysis – and is a clear gap in need of future work. For the present study, the statistics available allowed us to observe that, of the apps considered in this study, COVID Alert had the lowest download rate. The number of active users as a percentage of the population was also lower than for the comparison apps; however, the active users as a percentage of the number downloads was comparable to other apps.

**Table 1.**
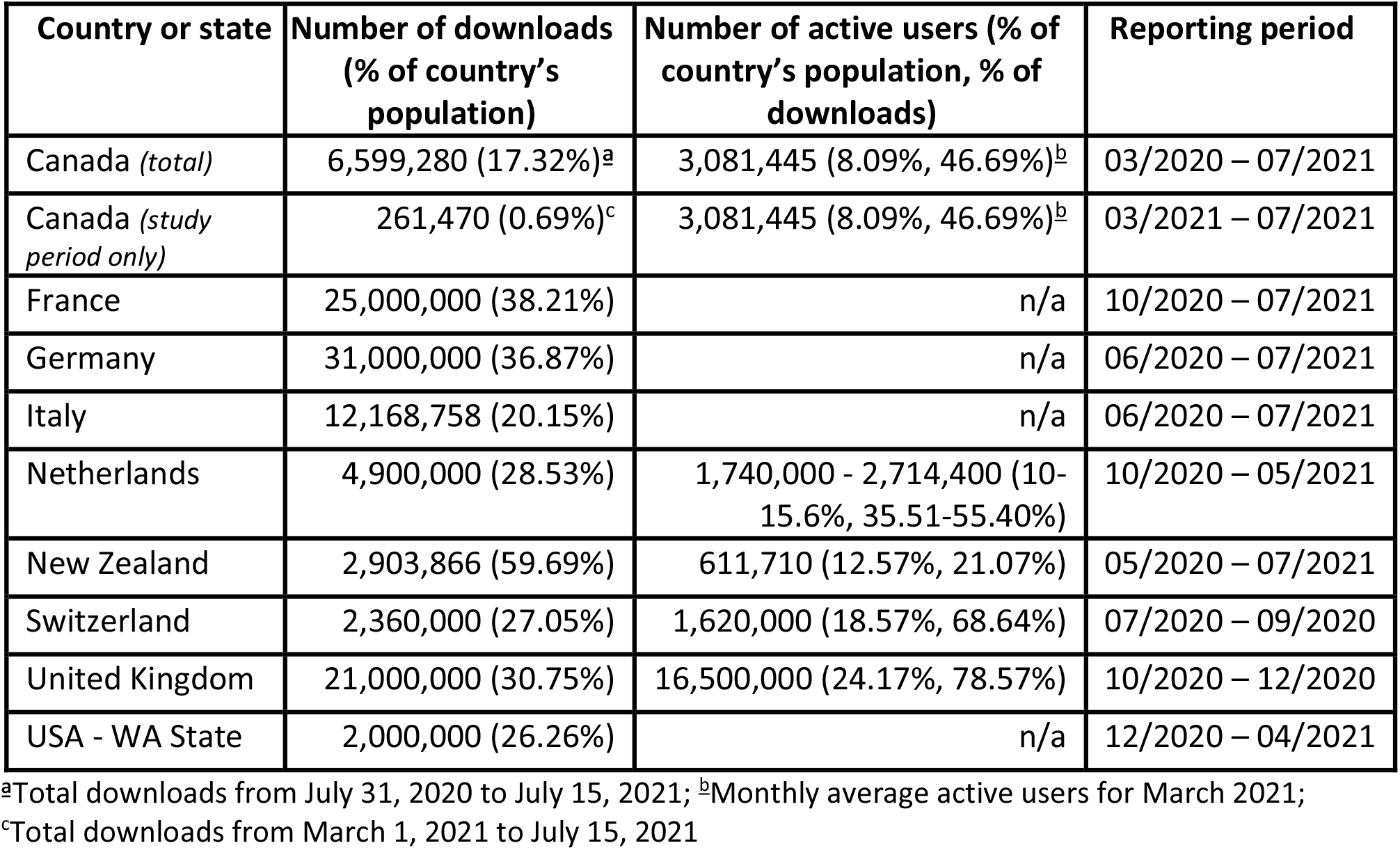
Total downloads and active users by country or state and the period of assessment for effectiveness reports.

| Country or state | Number of downloads<br>(% of country's<br>population) | Number of active users (% of<br>country's population, % of<br>downloads) | Reporting period |
| --- | --- | --- | --- |
| Canada (*total*) | 6,599,280 (17.32%)<sup>a</sup> | 3,081,445 (8.09%, 46.69%)<sup>b</sup> | 03/2020 – 07/2021 |
| Canada (*study*<br>*period only*) | 261,470 (0.69%)<sup>c</sup> | 3,081,445 (8.09%, 46.69%)<sup>b</sup> | 03/2021 – 07/2021 |
| France | 25,000,000 (38.21%) | n/a | 10/2020 – 07/2021 |
| Germany | 31,000,000 (36.87%) | n/a | 06/2020 – 07/2021 |
| Italy | 12,168,758 (20.15%) | n/a | 06/2020 – 07/2021 |
| Netherlands | 4,900,000 (28.53%) | 1,740,000 - 2,714,400 (10-15.6%, 35.51-55.40%) | 10/2020 – 05/2021 |
| New Zealand | 2,903,866 (59.69%) | 611,710 (12.57%, 21.07%) | 05/2020 – 07/2021 |
| Switzerland | 2,360,000 (27.05%) | 1,620,000 (18.57%, 68.64%) | 07/2020 – 09/2020 |
| United Kingdom | 21,000,000 (30.75%) | 16,500,000 (24.17%, 78.57%) | 10/2020 – 12/2020 |
| USA - WA State | 2,000,000 (26.26%) | n/a | 12/2020 – 04/2021 |

<sup>a</sup>Total downloads from July 31, 2020 to July 15, 2021; <sup>b</sup>Monthly average active users for March 2021;
<sup>c</sup>Total downloads from March 1, 2021 to July 15, 2021

### 3.2 Modeling

The estimated number of cases averted and the estimated number of deaths averted between 03/03/2021 and 15/07/2021 are listed in Table 2 and Table 3 for each province considered. We limited the assessment to include only provinces that sent >200 notifications during the assessment period: Manitoba, Newfoundland and Labrador, Nova Scotia, Ontario, Quebec, and Saskatchewan.

**Table 2.** Estimated total number of cases averted in select Canadian provinces between 03/03/2021 and 15/07/2021.

| Province | 45% quarantine effectiveness |  | 65% quarantine effectiveness |  |
| --- | --- | --- | --- | --- |
|  | 5% SAR | 6% SAR | 5% SAR | 6% SAR |
| Manitoba | 662 | 795 | 956 | 1,148 |
| Newfoundland and Labrador | 172 | 206 | 248 | 298 |
| Nova Scotia | 232 | 279 | 336 | 403 |
| Ontario | 4,567 | 5,481 | 6,597 | 7,917 |
| Quebec | 349 | 419 | 504 | 605 |
| Saskatchewan | 302 | 362 | 436 | 523 |
| **Total** | **6,284** | **7,542** | **9,077** | **10,894** |

**Table 3.** Estimated total number of deaths averted in select Canadian provinces between 03/03/2021 and 15/07/2021.

| Province | 45% quarantine effectiveness |  | 65% quarantine effectiveness |  |
| --- | --- | --- | --- | --- |
|  | 5% SAR | 6% SAR | 5% SAR | 6% SAR |
| Manitoba | 7 | 8 | 10 | 12 |
| Newfoundland and Labrador | 1 | 1 | 1 | 1 |
| Nova Scotia | 1 | 2 | 2 | 3 |
| Ontario | 42 | 51 | 61 | 74 |
| Quebec | 3 | 4 | 5 | 6 |
| Saskatchewan | 3 | 3 | 4 | 5 |
| **Total** | **57** | **69** | **83** | **101** |

Our analyses suggest that a considerable number of cases were averted by usage of the app. For the subset of provinces included in this assessment, the estimates range from approximately 6,284 to 10,894 cases averted, depending on the chosen parameters (Table 2). By multiplying each province-specific estimate of the number of cases averted by the province-specific crude case fatality rate observed for the same period, we estimate the number of deaths averted to be approximately 57 to 101, depending on the parameters chosen.

The number of cases averted is positively related to the severity of the pandemic (and the total number of notifications sent), with higher numbers of averted cases in areas with high numbers of confirmed cases (see Table 4). The results show that the ratio of cases averted to confirmed cases was, in general, higher in areas where a larger proportion of the population adopted the app. For example, in the more optimistic quarantine effectiveness scenario of 65% with the higher SAR of 6%, Newfoundland and Labrador, where the proportion of app users is 22.8%, had a higher ratio of cases averted to confirmed cases than Saskatchewan, where app usage was only 6.5%. This result suggests that a larger number of cases of COVID-19 were averted by exposure notification through the COVID Alert app in areas with a higher fraction of active app users in the population. Note that four components in the model, i.e., SAR, the expected fraction of transmissions prevented, the quarantine effectiveness and the expected fraction of transmissions prevented, are constant among the six selected provinces. Therefore Ontario, where the largest number of notifications were sent, has the greatest number of cases and deaths averted during this study period.

**Table 4.** Estimated total number of cases, notifications, proportion of app users, ratio of cases averted to cases, and ratio of deaths averted to deaths in select Canadian provinces between 03/03/2021 and 15/07/2021. Source for cases and notifications: CDS Github repository (Canadian Digital Service, n.d.).

| Province | Cases | Notifications<br>(% of app users) | Ratio of cases averted to cases |  | Ratio of deaths averted to deaths |  |
| --- | --- | --- | --- | --- | --- | --- |
|  |  |  | Lower bound | Upper bound | Lower bound | Upper bound |
| Manitoba | 25,078 | 5,622 (9.0%) | 0.026 | 0.046 | 0.026 | 0.046 |
| Newfoundland and Labrador | 439 | 564 (22.8%) | 0.392 | 0.679 | 0.392 | 0.679 |
| Nova Scotia | 4,227 | 1,288 (12.9%) | 0.055 | 0.095 | 0.055 | 0.095 |
| Ontario | 244,900 | 55,388 (9.7%) | 0.019 | 0.032 | 0.019 | 0.032 |
| Quebec | 87,168 | 4,138 (7.9%) | 0.004 | 0.007 | 0.004 | 0.007 |
| Saskatchewan | 20,403 | 3,170 (6.5%) | 0.015 | 0.026 | 0.015 | 0.026 |
| **Total** | **382,215** | **70,170 (9.4%)** | **0.016** | **0.029** | **0.016** | **0.029** |

Note: Lower bound: 5% SAR and 45% quarantine effectiveness; Upper bound: 6% SAR and 65% quarantine effectiveness.

Figure 1 displays the upper- and lower-bound estimates of the total number of daily cases averted by the COVID Alert app in the six provinces, along with the total number of observed daily cases during this study period. For a more detailed picture, please see Web Figure 1, which illustrates the upper-bound estimates of daily cases averted by province over the study period. The shape of estimated cases averted captures the trend of daily confirmed cases, and Ontario has the greatest number of cases averted.

**Figure 1.**
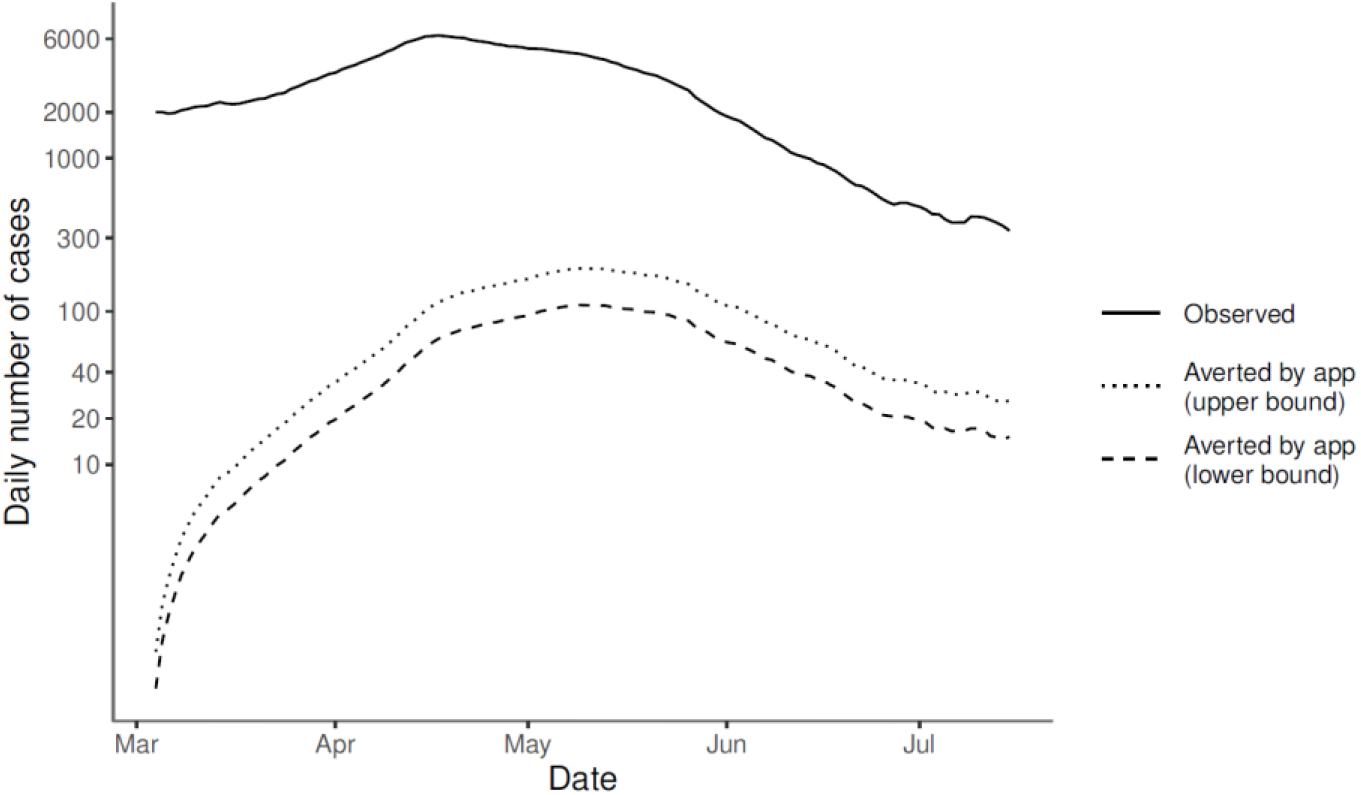
Estimated daily COVID-19 cases averted over time.

Estimated cases averted, estimated deaths averted, and exposure notifications for COVID Alert are summarized in Tables 2-4. Based on the available literature, we also provide comparisons with other notification apps (Table 5). Data on estimated efficacy information was scarce; only three of eight countries provided such estimates, and four of eight had exposure notification data available. Given the available data, our analyses suggest comparable efficacy between COVID Alert and the apps from Italy, France, and Washington state.

**Table 5.** Reported app effectiveness for a subset of the regions with COVID notification apps.

| Country or state | Total exposure notifications (% of total cases) | Estimated cases averted (% of total cases) | Estimated deaths averted (% of total cases) |
| --- | --- | --- | --- |
| Canada | 74,562 (5.18 - 5.22%) | 6,284 -10,894 (0.47-0.81%) | 57-101 (0.004 - 0.01%) |
| France | 200,037 (3.32%) | n/a | n/a |
| Italy | 101,051 (2.34%) | n/a | n/a |
| Netherlands | n/a | 45,088 (2.44%) | 271 (0.01%) |
| United Kingdom | 1,700,000 (29.59%) | 284,000-594,000 (4.94-10.34%) | 4,100-8,700 (0.07-0.15%) |
| USA - WA state | 34,501 (7.32%) | 8,547 (1.81%) | 40-115 (0.01-0.02%) |

Notes: 1) Total cases for Canada includes estimated cases averted. 2) The number of notifications listed here is for the whole of Canada and not simply the six provinces listed in Table 4.

## 4. Discussion

The analyses presented here demonstrate that the adoption of the COVID Alert app in Canada contributed to mitigating the COVID-19 pandemic. While our analysis was dependent on some modelling parameters drawn from other countries, we estimated that the COVID Alert app could have averted up to (i.e., upper bounds on estimates were equal to) 10,894 cases and 101 deaths in the six provinces where usage was highest between 03/03/2021 and 15/07/2021. Most notably, the results indicate that the proportion of app users is positively associated with the ratio of cases averted to cases, which supports the positive intervention effects of COVID Alert on reducing the burden of the disease. The number of cases and deaths averted was largest in Ontario, however the impact in terms of the proportion of cases and death averted was greatest in Newfoundland and Labrador.

There are several key observations that may be taken from our analysis. First, when adopted widely, the COVID Alert app averted appreciable numbers of cases and deaths. Newfoundland and Labrador as well as Nova Scotia had relatively large app installation and usage, which translated into a high proportion of averted infections and deaths in these provinces. In contrast, provinces with lower adoption of the app had much lower estimates of the number of averted infections and deaths. Notably, this association is not simply a product of our parameter assumptions but is reflective of the underlying data, specifically the ratio of exposure notifications to population size and province-specific case fatality rates. This provides a strong indication that the COVID Alert app has the capacity to be an effective tool for mitigating infections and deaths due to COVID-19. However, this efficacy (much like vaccines and other public health instruments) is dependent on widespread adoption.

Nationally, the COVID Alert app had limited impact. Because the app was not widely adopted in some of the more populated provinces, its overall impact was quite limited, preventing only 1.6-2.9% of cases and deaths during the study period. Among similar exposure notification apps that have reported data, COVID Alert had among the lowest adoption rates. However, attempting to compare the adoption of the app against other apps is fraught with difficulty due to differences in the reporting time periods. While adoption rates *among countries that reported this for their apps* were generally low (20-38%) with the exception of New Zealand at nearly 60%, adoption of the COVID Alert app was the lowest at about 17.3%.

An important limitation of our comparative analysis is that there were insufficient data available to provide reliable comparisons of the relative impact of exposure notification apps across countries. Because app deployment analyses and reporting periods differ across time (see Table 1), any direct comparisons across countries are not well defined, particularly given the fluctuating nature of COVID cases or “waves.” Reports span varying time periods, and it is not possible to assess if a given report provides accurate or representative overall usage and efficacy of an app or whether instead it captures a period of particular efficacy or lack thereof. Additionally, different apps had different purposes and resources dedicated to their development. For example, COVID Alert was developed and used as an exposure notification app, while the NHS COVID-19 App had additional functionality (e.g., checking local alert level, checking symptoms, booking COVID-19 tests) in addition to exposure notification. The paucity of data currently available from other countries makes meaningful comparisons of efficacy impossible. Further, given different app purposes and associated budgets, combined with the scarcity of published or public data, comparing costs of development and deployment was not possible.

The main limitation of our modelling analysis is the inability to estimate key parameters of the model due to the high level of data aggregation employed to preserve privacy. Rather than being estimated directly from Canada-specific data, these parameters had to be assumed and were chosen based on the literature. Thus, the reliability of our findings is dependent on the fidelity of our assumptions to the truth, which is empirically untestable with the available data. In particular, the values of the effectiveness of quarantine were estimated based on surveys in the UK and the USA, but it is difficult to assess their reliability and comparability to the effectiveness of quarantine in Canada. Furthermore, the fraction of the transmission chain prevented by notification depends on the time elapsed between exposure and notification, which could vary across the time period of the analysis. For the aggregate data available to us, we used a single estimated value for this study period, i.e., 5.46 days between exposure and notification. For the period of assessment considered here, this assumption that there is no variation in elapsed time is not unreasonable as delay times for testing and reporting were generally fairly uniform by March 2021.

A second important limitation of this modelling is that exposure notification data were unavailable prior to the last few days of February 2021. We were thus unable to provide impact estimates for the entire time period during which the app was used (piloted first in Ontario in July 2020 and then deployed more widely in the beginning September 2020). Given the dramatic variation in infection rates and changes in population behaviour as waves hit different provinces, as well as the variety of measures taken by individual local and provincial governments in response, no attempt was made to extrapolate to time periods prior to March 2021. Nonetheless, trends in the number of downloads provides a strong indication that adoption of the app was higher in the months *before* February 2021. This almost certainly would correlate with more active users, higher clustering of users within the population, and, as a result, more exposure notifications per case. Accordingly, it is highly probable that the true percent of cases and deaths averted since the launch of the app is higher than the estimates reported here, particularly given the large wave of cases that occurred in December 2020 - January 2021 across most of the country.

Finally, it must be noted that the majority of the data available came from the province of Ontario, the most populous province in the country. As our analyses were province-specific, this did not unduly influence the estimated proportion of cases and deaths averted in other regions of the country, however when aggregating the total numbers averted, Ontario data dominate.

This analysis invites reflection on the data collected and how it impacted the modeling that one might like to perform in contrast with that which could be done. A key observation is that purposefully designed data collection with a specific modeling technique in mind could assist in understanding (and improving) the efficacy of a notification app. The data available, while useful, were incomplete for the modeling performed here and, most likely, for any impact-oriented modeling task. The COVID Alert app was widely hailed for its privacy (Daigle, 2020; Davis, 2020). It is possible that had impact modeling been included as another design consideration at the outset, the data needed would have been collected, and the data that were belatedly collected (such as the number of notifications) would have been available for a much longer period without compromising privacy guarantees.

Overall, our findings give support to the use of exposure notification apps as tools in the mitigation and management of epidemic events. For such apps to be useful, however, they must be sufficiently adopted and protocols around their usage implemented. The way in which these factors diminished the ultimate impact of the COVID Alert app underscores their importance for future public health research. Unfortunately, the feasibility of a widespread adoption of such an exposure notification app is unclear. With healthcare provincially controlled, the promotion and consequent uptake of such an app may vary considerably across the country and the cost-benefit assessment of public health versus economic impacts are not agreed upon – particularly in settings where the cost of the app may be borne federally while its benefits more clearly observed in terms of reduction in provincial healthcare expenditures. Further, the size of the epidemic can change usage and consequent impact. For instance, based on the data presented in our comparative analysis, the UK’s exposure notification app *appears* to be a model to be emulated, as it saw high uptake, averted many cases and deaths, and collected sufficient data to estimate many of the parameters needed to understand its impact. However, following a general easing of restrictions, the UK experienced a surge in cases and, in consequence, what was referred to as a “pingdemic” in which large numbers of people were required to isolate following notification of an exposure -- more than half a million in England just in the first week of July 2021 (Rimmer, 2021). These high rates of notification were seen by many as having a detrimental effect on industry and led some employers to recommend uninstalling the app (Lawton, 2021). Thus, much remains to be understood about the sociological, societal, and economic impact of exposure notification apps under varying conditions of outbreak and spread.

## Data Availability

All data produced in the present study are available upon reasonable request to Health Canada at.

## Appendix

In this Appendix, we provide detailed definitions of the terms needed to model the impact of the exposure notification app on averted cases due to COVID-19.

### The number of notifications received

The process to obtain a one-time key to upload a positive COVID-19 test is different in each province and territory, thus it is possible that some users do not ever receive a key. Further, COVID-19 positive declaration is not mandatory and users who test positive have only 24 hours to enter the key in the app. Therefore, the number of notifications received during the study period is an underestimate of the number of users who test positive.

### Secondary attack rate

The secondary attack rate (SAR) is defined as the probability that an infection occurs among individuals notified by the COVID Alert app within a reasonable incubation period (e.g., 14 days). It can provide an indication of how social interactions relate to transmission risk. The SAR in Wymant et al. (2021) is 6.02% (CI: 5.96 - 6.09%); the lower bound of SAR estimate in Segal et al. (2021) is 5.1%. We therefore consider two plausible SAR values: 5% and 6%.

### The generation time distribution

The generation time (or generation interval) is defined for source-recipient transmission pairs as the interval between the time of infection of the index case and the time of infection of the secondary case. It is typically difficult to estimate this quantity directly unless the interval of exposure is short for both the source case and the recipient case. Instead, the generation time is usually estimated indirectly from intervals of exposure and onset of symptoms. Ferretti et al. (2020) directly estimate the generation time distribution from 191 source-recipient pairs, with known time of onset of symptoms, intervals of exposure, and a meta-distribution of incubation period. The distribution is described by a Weibull distribution (shape=3.2862 and scale=6.1244) with mean generation time equal to 5.5 days (Ferretti et al. 2020b).

### The expected fraction of transmissions prevented by strict quarantine

The expected fraction of transmission preventable if an infectious individual strictly adheres to quarantine after receiving a notification depends on the delay time, i.e., the interval (delay) between exposure and exposure notification. We adopt the estimated mean time of 5.46 days from Segal et al. (2021). The expected fraction of transmissions prevented by receipt of exposure notification is the upper tail probability of the generation time distribution (i.e., P(X ≥ 5.46)), which corresponds to approximately 0.50.

### The quarantine effectiveness

Adherence to quarantine is critical but difficult to access reliably. The plausible values, 45% and 65%, are considered based on estimations in Wymant et al. (2021) and Segal et al. (2021) in which effectiveness of quarantine is estimated based on surveys and assumptions. The overall effectiveness of quarantine estimation is 61% (with range from 53.25% to 68.74%) in Wymant et al. (2021), and the estimate in Segal et al. (2021) is 53%.

### The expected size of the full transmission chain

The size of the transmission chain is a function of the number of cases during the study period. Let *d*(*x, t*) denote the number of diagnosed cases among the whole population in province *x* at time *t*. The number of cases averted at time *T* (i.e., the size of the whole chain caused by a single transmission at time *t*), is 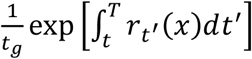 for any *t* < *T*, where *t*_*g*_ is the mean generation time and *r*_*t*_(*x*) is the epidemic growth rate at time *t*. Here we follow the assumptions of Wymant et al., 2021; specifically, it is assumed that local epidemics do not mix and that the extra cases do not affect the epidemic dynamic (i.e., the underlying growth rate *r*_*t*_(*x*) does not change with the additional cases). Then we obtain 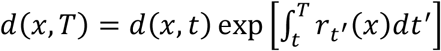 for any *t* < *T*. Therefore, instead of estimating the growth rate *r*_*t*_(*x*), we can use the factor 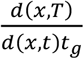 to obtain the number of cases averted at time *T* for each transmission time *t*. The number of cases *d*(*x, t*) is estimated by taking the weekly moving average of new daily cases in each Canadian province.

**Appendix Figure 1.**
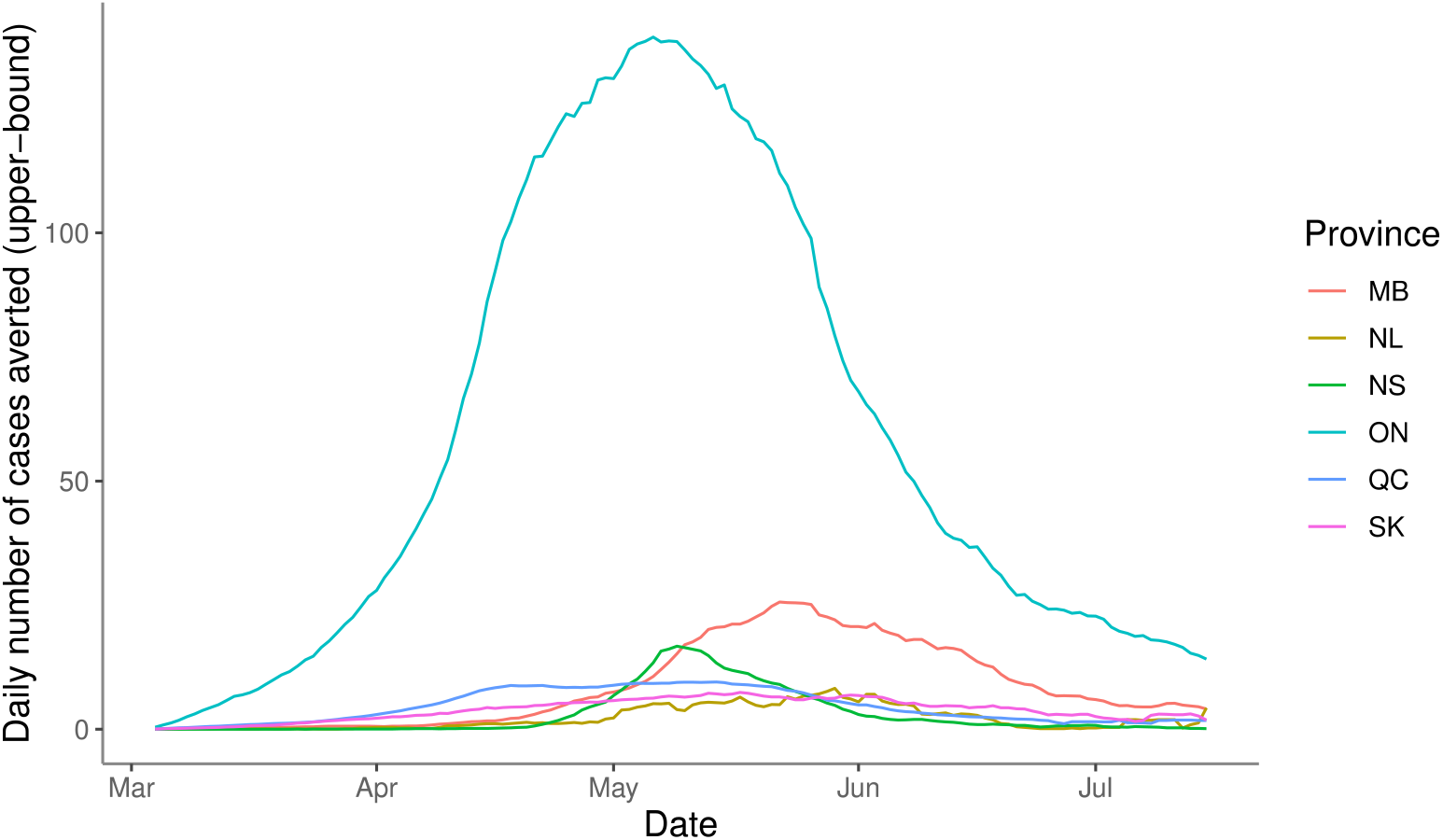
Estimated daily COVID-19 cases averted by province over time.

